# Childhood Immuno-metabolic Markers and Risk of Depression and Psychosis in Adulthood: A Prospective Birth Cohort Study

**DOI:** 10.1101/2021.11.19.21266562

**Authors:** NA Donnelly, BI Perry, HJ Jones, GM Khandaker

## Abstract

**Background:** Metabolic and inflammatory disorders commonly co-occur with depression and psychosis, with emerging evidence implicating immuno-metabolic dysfunction in their aetiology. Previous studies have reported metabolic dysfunction and inflammation in adults with depression and psychosis. However, longitudinal studies testing the direction of association, and the effects of different dimensions of early-life immuno-metabolic dysfunction on adult psychopathology, are limited.

**Methods:** Using data from 3875 birth cohort participants we examined longitudinal associations of three metabolic hormones (leptin, adiponectin, insulin) at age 9 with risks for depression- and psychosis-spectrum outcomes at age 24. In addition, using nine immuno-metabolic biomarkers, we constructed an exploratory bifactor model showing a general immuno-metabolic factor and three specific factors (adiposity, inflammation, and insulin resistance), which were also used as exposures.

**Results:** Childhood leptin was associated with adult depressive episode (adjusted odds ratio (aOR)=1.28; 95% CI, 1.00-1.64) and negative symptoms (aOR=1.12; 95% CI, 1.05-1.20). The general immuno-metabolic factor was associated with depressive symptoms (aOR=1.05; 95% CI, 1.01-1.08) and psychotic experiences (aOR=1.20; 95% CI, 1.01-1.42). The adiposity factor was associated with negative symptoms (aOR=1.07; 95% CI 1.02-1.12). All associations tended to be stronger in women, though 95% credible intervals overlapped with that for men. In women, the inflammatory factor was associated with depressive episode (aOR=1.23; 95% CI, 1.01-1.47) and atypical depressive symptoms (aOR=1.10; 95% CI, 1.02-1.19).

**Conclusions:** While general immuno-metabolic dysfunction in childhood may contribute to risks for both psychotic and depressive symptoms in adulthood, childhood adiposity and inflammation are linked to affective (depressive, atypical, and negative) symptoms.

## 1. Introduction

Immuno-metabolic alterations are present in depressive and psychotic disorders, and are associated with excess mortality (Hoang et al., 2013; Milaneschi et al., 2020, 2019; Vancampfort et al., 2015). The prevalence of cardiometabolic disease (Penninx et al., 2001; van Melle et al., 2004), autoimmune inflammatory conditions (Benros et al., 2011; Eaton et al., 2006), and levels of circulating inflammatory markers (Khandaker et al., 2014; Osimo et al., 2019), are higher in people with depression and psychosis compared with controls. However, the direction of these relationships remains unclear. For example, while psychotropic medications can increase predisposition to cardiometabolic disorders (Vancampfort et al., 2015), there is evidence that predisposition towards cardiometabolic dysfunction predates treatment with psychotropic medications (Benseñor et al., 2012; Perry et al., 2016, 2021b; Pillinger et al., 2017). Therefore, longitudinal studies are required to address the issue of direction of association.

Emerging evidence from longitudinal studies suggests that inflammation may predate the onset of depression and psychosis. Using longitudinal data from the Avon Longitudinal Study of Parents and Children (ALSPAC) and Northern Finland Birth Cohort (NFBC) 1986 birth cohorts (Khandaker et al., 2014; Metcalf et al., 2017; Perry et al., 2021b), we have reported associations between higher circulating levels of interleukin 6 (IL-6) and C-reactive protein (CRP) in childhood and adolescence with risk of depression and psychosis subsequently in adulthood. Similar longitudinal associations have also been reported from the UK Whitehall (Gimeno et al., 2009), Dutch Generation R (Zalli et al., 2016) and the Netherlands Study of Depression and Anxiety (NESDA) (Lamers et al., 2020) cohorts. Using longitudinal repeated measures data from the ALSPAC cohort, we have reported higher risk of psychosis in adulthood associated with persistently high insulin levels across childhood, adolescence and early-adulthood, which may vary by sex (Perry et al., 2021b). These findings point to a role of immuno-metabolic alterations in childhood, a critical period for the development of the nervous, immune, and endocrine systems, in the aetiology of psychiatric disorders.

Despite previous studies reporting obesity, altered glucose-insulin homeostasis, lipids, and other metabolic dysfunction in people with depression, psychosis and young people with psychotic experiences (Milaneschi et al., 2019; Perry et al., 2019; Vancampfort et al., 2015), longitudinal studies of childhood metabolic markers and subsequent depression and psychosis are relatively scarce. Particularly lacking are studies of metabolic hormones such as insulin, adiponectin and leptin, which are key neuroendocrine regulators of energy homeostasis. Immuno-metabolic biomarkers are interlinked and have pleiotropic functions. For instance, obesity is associated with systemic low-grade inflammation (Timpson et al., 2011; Yudkin et al., 1999), while insulin and leptin are influenced by inflammation and obesity (Considine et al., 1996; Jerry and Lesley, 2006; van Dielen et al., 2001). Commonly known as a satiety hormone, leptin is structurally similar to IL-6 and may have inflammatory functions (Abella et al., 2017). Additionally, previous studies have typically considered only a few immune or metabolic markers in isolation and ignored the underlying structure of covariance between biomarkers.

To address the issue of direction of association, we have carried out a longitudinal study testing the associations of three key metabolic hormones (insulin, leptin and adiponectin), measured in childhood at age 9 years, with several outcomes across the spectrum of depression (depressive episode, total symptom score, and atypical symptom score) and psychosis (psychotic disorder, at-risk mental state, psychotic experiences, negative symptoms) at age 24 years, in a whole analysis sample and in men and women separately, using data from the ALSPAC birth cohort. Furthermore, combining these hormones with related immuno-metabolic markers (BMI, LDL, HDL, triglycerides, IL-6, and CRP), also measured at age 9, we identified factors representing general and specific dimensions of early-life immuno-metabolic dysfunction, and tested their associations with psychiatric outcomes at age 24.

## 2. Materials and Methods

### 2.1 Cohort Profile

We used data from the ALSPAC birth cohort (Boyd et al., 2013; Fraser et al., 2013; Northstone et al., 2019), which recruited 14,541 pregnant women in the Avon area of the United Kingdom between 1990 and 1992; of these initial pregnancies 13,988 children were alive at 1 year of age. Further phases of recruitment occurred when the oldest children in the cohort were approximately 7 years of age, resulting in an additional 913 children being enrolled; giving a total cohort size of 15,454 pregnancies, resulting in 15,589 deliveries, of which 14,901 were alive at 1 year of age. The study is considered to be representative of the UK population (Golding et al., 2001).

Study data are collected and managed using the REDCap (Research Electronic Data Capture) tools hosted at the University of Bristol (Harris et al., 2019, 2009). Children and parents have been subsequently followed up with clinic and questionnaire assessments for a range of health outcomes. The ALSPAC study website contains details of all available data through a fully searchable data dictionary http://www.bristol.ac.uk/alspac/researchers/our-data/.

Ethical approval for the study was obtained from the ALSPAC Ethics and Law Committee and the Local Research Ethics Committees (http://www.bristol.ac.uk/alspac/researchers/research-ethics/). Consent for biological samples was collected in accordance with the UK Human Tissue Act (2004). Informed consent for the use of data collected via questionnaires and clinics was obtained from participants following the recommendations of the ALSPAC Ethics and Law Committee at the time.

This study is reported in line with the Strengthening the Reporting of Observational Studies in Epidemiology (STROBE) reporting guideline for cohort studies.

### 2.2 Measurement of childhood immuno-metabolic biomarkers at age 9

#### 2.2.1 Metabolic Hormones

Leptin, adiponectin and random insulin were measured at the cohort age 9 clinic visit (**Supplementary Table 1**). As previously described (Falaschetti et al., 2010), non-fasting blood samples were collected during clinic visits, using standard aseptic procedure and immediately spun and frozen for long-term storage at -80°C. Blood biomarkers were assayed in 2008 after a median of 7.5 years in storage with no previous freeze–thaw cycles during storage. We did not include fasting insulin in this study due to the lower sample size available for that measure (Ong et al., 2004). Leptin (ng/ml) was measured by an in-house enzyme-linked immunosorbent assay (ELISA) validated against commercial methods. Adiponectin (ng/ml) and random insulin (mU/l) was measured using an ultrasensitive automated microparticle enzyme immunoassay (Mercodia), which does not cross-react with proinsulin. All assay coefficients of variation were <6%.

#### 2.2.2 Other Metabolic Biomarkers

We included plasma lipids (LDL, HDL and triglycerides (mmol/l)), also measured at the cohort age 9 clinic visit (**Supplementary Table 1**). All were measured by a modification of the standard Lipid Research Clinics Protocol using enzymatic reagents for lipid determination.

#### 2.2.3 Inflammatory Biomarkers

We included IL-6 and CRP, which were also measured at the cohort age 9 clinic visit (**Supplementary Table 1**). IL-6 (pg/ml) was measured by ELISA (R&D systems, Abingdon, UK), and CRP (mg/l) was measured by automated particle-enhanced immunoturbidimetric assay (Roche UK, Welwyn Garden City, UK).

### 2.3 Assessment of Psychiatric Outcomes at age 24

#### 2.3.1 Psychosis-Spectrum Outcomes

Psychotic positive symptoms were measured at age 24 with the psychosis-like symptoms (PLIKS) interview (Sullivan et al., 2020; Zammit et al., 2013). Interviews were undertaken using a semi-structured format by trained psychologists. Psychotic experiences (PEs) were defined as symptoms in 3 domains (hallucinations, delusions, and thought interference) occurring in the 6 months prior to the interview. After cross-questioning, PEs were rated as absent, suspected, or definite. We included definite PEs as our outcome of interest.

Psychotic disorder was defined as definite PLIKS, not attributable to sleep or fever, which occurred at least once a month in the past 6 months, which either caused the respondent to seek professional help, was very upsetting/distressing, and/or had a negative impact on their social/occupational life.

Cases of At Risk Mental State (ARMS) were defined by mapping the Comprehensive Assessment of the At Risk Mental State (CAARMS) criteria onto the PLIKS variables (Perry et al., 2021b). ARMs were defined as participants meeting CAARMS criteria for attenuated psychosis (symptoms not reaching the psychosis threshold owing to levels of intensity or frequency), brief limited intermittent psychosis (frank psychotic symptoms that resolved spontaneously within 1 week), or the CAARMS psychosis group.

Negative symptoms at age 24 were measured using ten questions from a self-administered, computerised version of the Community Assessment of Psychic Experiences (CAPE) questionnaire (Jones et al., 2016; Konings et al., 2006; Stefanis et al., 2002). We generated a single outcome variable by dichotomising responses to each question (each item was rated by participants on a 1-4 scale, we grouped responses 1 and 2 as “symptom present”, scoring 1 and responses 3 and 4 as “symptom absent”, scoring 0) and summing the resultant scores, giving each participant a negative symptom score from 0 - 10.

#### 2.3.2 Depression Outcomes

Depressive symptoms at age 24 were measured using a self-administered, computerised version of the Clinical Interview Schedule, Revised (CIS-R) (Lewis et al., 1992). We took three outcomes from the CIS-R data: (1) a dichotomous depressive episode variable, based on a mapping of the CIS-R responses onto ICD-10 depression diagnostic criteria; (2) a symptom score which was constructed as a sum of scores of the mood, thoughts, fatigue, concentration, and sleep components for the CIS-R. This score ranged between 0 and 20. We also derived (3): an atypical depression symptoms score composed of the sum of participants scores on 4 items from the CIS-R: increased appetite, increased weight, hypersomnia and low energy. Note that the CIS-R does not contain a measure of “leaden paralysis” as has been included in other measures of atypical depression (Lamers et al., 2020), as this measure is not part of many common depression screening instruments (Fried, 2017).

#### 2.3.3 Confounders

We included maternal social class (determined using occupational status) and maternal highest educational level as potential confounders. IL-6 and BMI at age 9 (weight in kg divided by height in meters squared) were also included for analyses where they were not part of the predictors.

### 2.4 Statistical Analysis

#### 2.4.1 Data Transformation

We log_10_-transformed all metabolic measures to give approximately normally distributed data, except for adiponectin, which was already approximately normally distributed. All continuous exposure and covariate variables were centred and standardised (mean value subtracted and then divided by the standard deviation). Therefore, odds ratios (ORs) represent change in outcome per SD change in exposure.

#### 2.4.2 Exploratory Factor Analysis

Using adiponectin, insulin, leptin, IL-6, CRP, HDL, LDL, triglycerides, and BMI, we explored the underlying factor structure of the immuno-metabolic phenotype at age 9. Before analysis, we inverted HDL to match the cardiometabolic risk-increasing direction of other markers.

Factor analysis was done using the exploratory factor analysis (EFA) function in the *EFAtools* R package. The number of factors was selected after inspecting the results of a principal components analysis (**Supplementary Methods, Supplementary Figure 2**). We fit a one factor, three factor, and a bifactor model with 3 subfactors, and compared model fit (**Supplementary Table 2**). Compared to a one- or 3-factor model, the bifactor model was a better fit to our data. We then extracted factor scores for each factor for each subject (**see Supplementary Figure 2 and Supplementary Table 3 for factor loadings**).

#### 2.4.3 Modelling Approach

We analysed data using R version 4.1.0 (R Development Core Team, 2017), fitting Bayesian regression models using the *brms* (Bürkner, 2017) and *rstanarm* packages (Carpenter et al., 2017). As the number of participants providing data for different outcomes varied, we fit each model to the largest possible dataset of complete cases for all covariates, exposures and outcomes (**see Supplementary Tables 4-6 for full details**).

We fit unadjusted models with a psychiatric outcome predicted by our exposure variables of interest (leptin, adiponectin and insulin). Next, we fit adjusted models with BMI at age 9, log_10_ IL-6, sex at birth and maternal socioeconomic class included as covariates. We also fit adjusted sex-stratified models to all female and male participants separately. For models using factor scores as exposures, we included all factor scores as exposures plus sex at birth and maternal socio-economic class as covariates.

We used regularising priors for all models (McElreath, 2018), calculating model estimate and the 95% credible interval, using the median and the highest density interval method. We present as summary statistics the probability of direction, which can be considered analogous to the frequentist p-value (Makowski et al., 2019a, 2019b), and the Region of Practical Equivalence (ROPE), the proportion of the posterior falling within a set region around the null, here defined as an odds ratio in the range 0.9 – 1.1 (Kruschke and Liddell, 2018). **See supplementary methods for further details**.

#### 2.4.4 Missing data

We analysed missingness in the dataset with two models: a model for missingness of clinic data at age 9, with predictors of maternal education, maternal social class and ethnicity and sex of the child; and a model for missingness for age 24 psychiatric data in the set of participants who had blood tests at age 9, with predictors of maternal education, maternal social class, ethnicity and sex of the child, and age 9 adiponectin, random insulin and leptin, IL-6 and BMI.

#### 2.4.5 Data Availability

In line with the ALSPAC data access policy, all raw data used for the work presented can be accessed by other investigators by making a request to the ALSPAC study executive (see http://www.bristol.ac.uk/alspac/researchers/access/).

## 3. Results

### 3.1 Childhood immuno-metabolic biomarker distributions, correlates, and factor score calculation

Data on three metabolic hormones (insulin, leptin, adiponectin) and six related immuno-metabolic biomarkers (BMI, HDL, LDL, triglycerides, IL-6, and CRP) were available from total 3,875 participants at age 9 (**Supplementary Table 1**).

Higher leptin was associated with higher BMI (beta = 0.71; 95% credible interval [CI] 0.69-0.73), female sex (beta for girls compared to boys = 0.48; 95% CI 0.44 - 0.51) and higher IL-6 (beta = 0.04; 95% CI 0.02-0.05). Higher adiponectin was associated with lower BMI (beta = -0.14; 95% CI -0.16 - -0.11), female sex (beta = 0.12; 95% CI 0.07-0.18) and higher IL-6 (beta = 0.04; 95% CI 0.01-0.06). Higher insulin was associated with higher BMI (beta = 0.22; 95% CI 0.19-0.25, **see Supplementary Figure 1 and Supplementary Table 7**).

Immuno-metabolic biomarkers were correlated (correlations ranged from -0.18 for adiponectin-HDL to 0.75 for leptin-BMI, **Figure 1**). Using data for nine biomarkers we calculated a bifactor model identifying a general immuno-metabolic factor (*G*, loading on all biomarkers), and three sub-factors representing: (1) adiposity (loading on higher BMI, leptin and LDL); (2) inflammation (loading on higher IL-6 and CRP); and (3) insulin resistance (loading on higher triglycerides, insulin and lower HDL, **Supplementary Table 3**). There was no relationship between sex or maternal class and *G;* all sub-factors had higher mean values in females (**Supplementary Table 8**).

**Figure 1:**
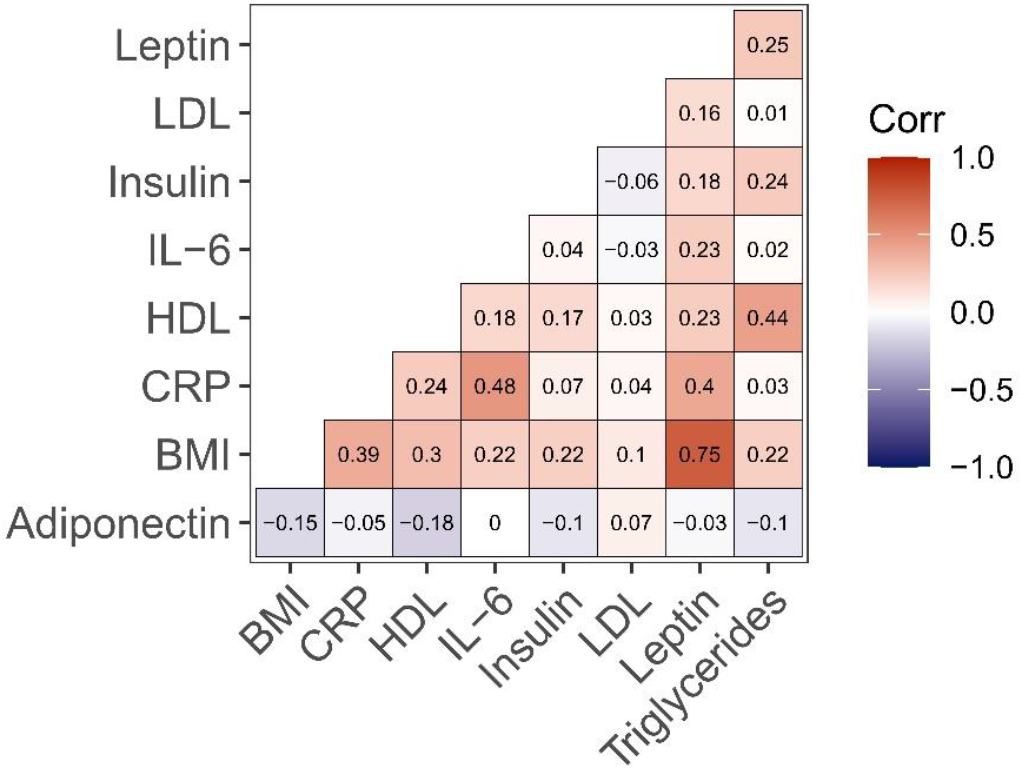
Correlation between immuno-metabolic biomarkers at age 9 in the ALSPAC birth cohort. Matrix of correlations between all 9 immuno-metabolic markers measured at age 9. Colour represents correlation; negative correlations are represented in blue, positive correlations in red. Numerical values are correlation coefficients.

### 3.2 Associations of childhood metabolic hormones with depression and psychosis risk in adulthood

#### 3.2.1 Whole Sample Analysis

After adjusting for potential confounders, leptin at age 9 was associated with depressive episode (adjusted odds ratio [aOR] =1.28; 95% CI, 1.00-1.64) and negative symptoms at age 24 (aOR=1.12; 95% CI 1.05-1.20). Adiponectin was also associated with negative symptoms (aOR=1.05; 95% CI 1.00-1.09 **Figure 2, Table 1**). Insulin was not associated with any psychiatric outcome in our total sample.

**Figure 2:**
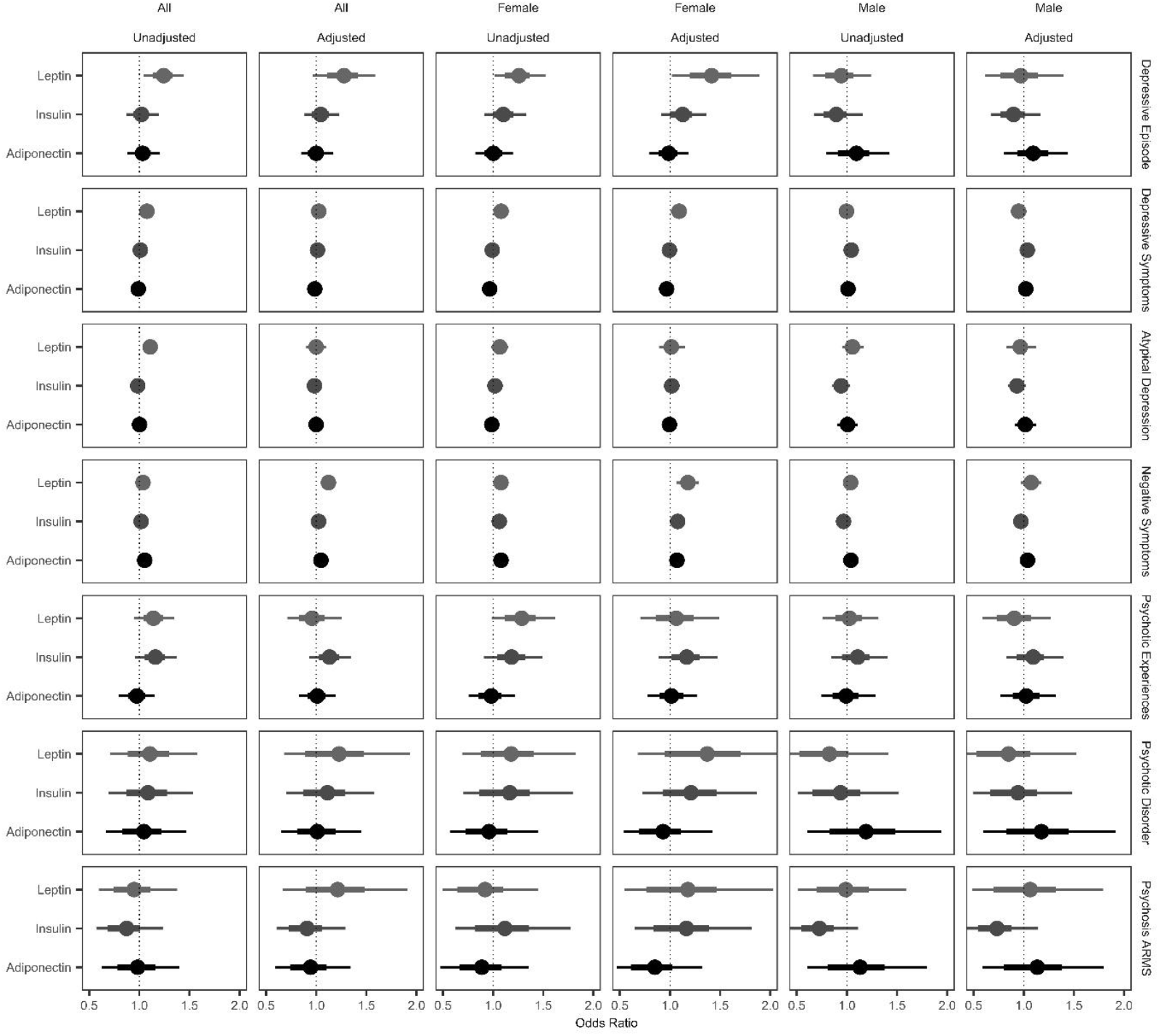
Associations of Childhood Metabolic Hormones with Adult Depression and Psychosis Measures in the Whole Sample. Plots of estimated odds ratios for the effect of our exposures (adiponectin, insulin and leptin) on psychiatric outcomes at age 24. Data are presented unadjusted and adjusted (for sex in the all-participant model, IL-6 at age 9, BMI at age 9 and maternal social class) and grouped by model: all participants, only females or only males. Thin bars represent the 95% credible interval, thicker bars the 66% credible interval, and dots the median of the posterior distribution for that parameter.

**Table 1:**
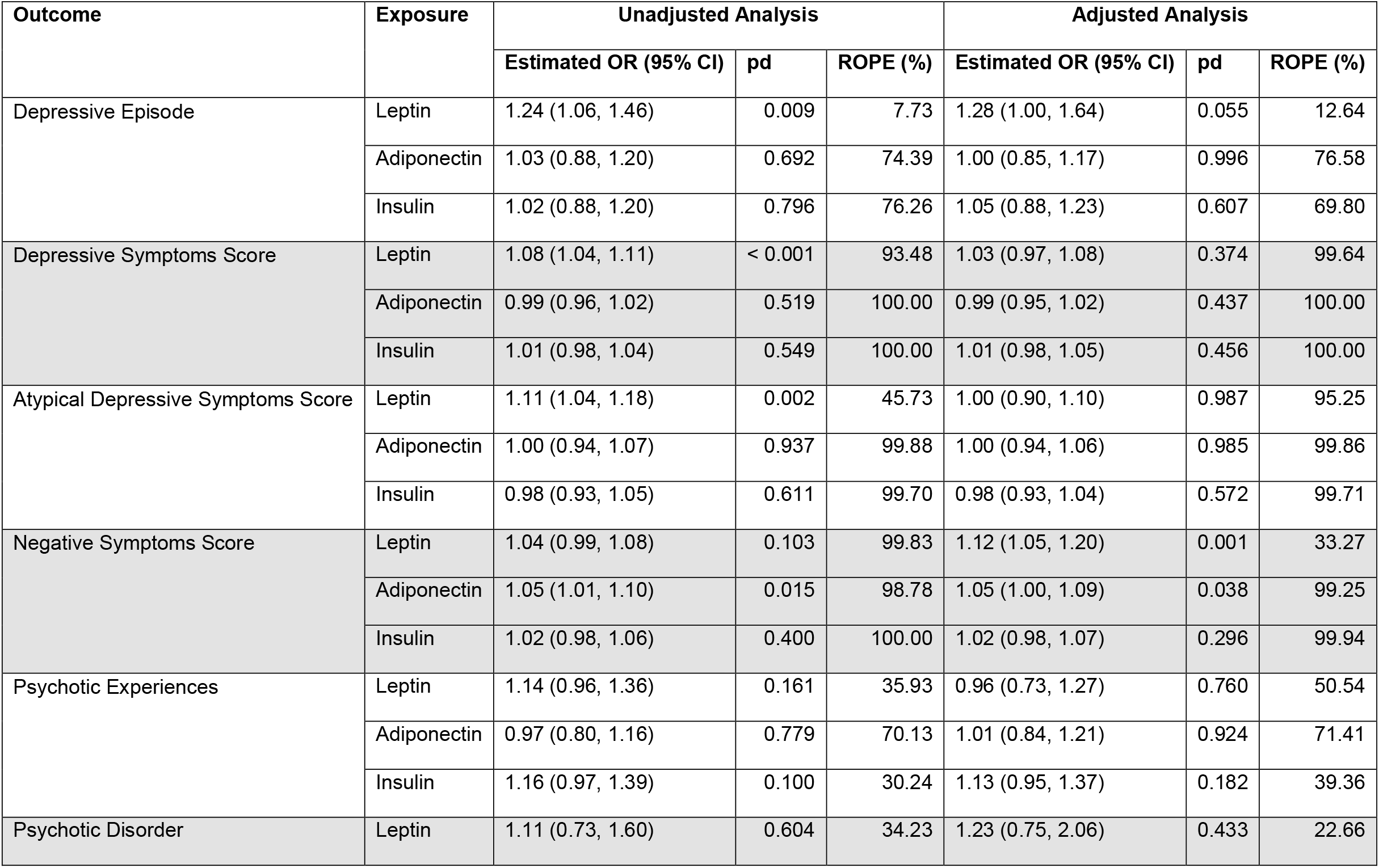
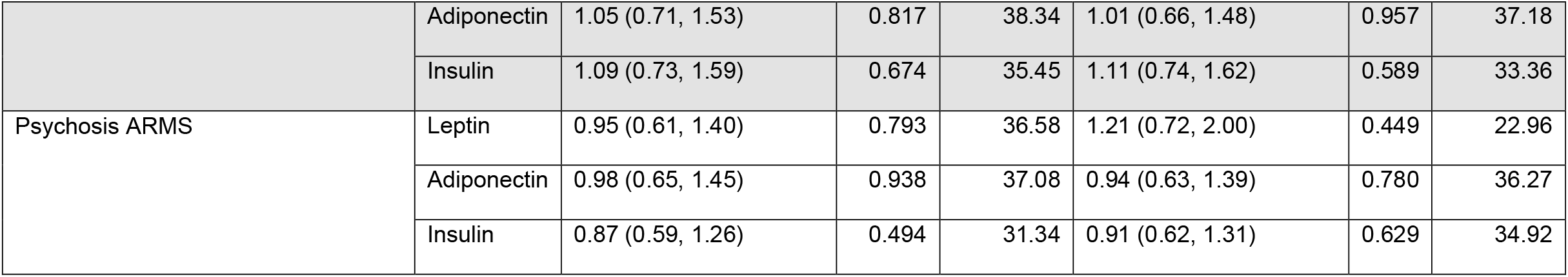
Associations of Childhood Metabolic Hormones with Adult Depression and Psychosis Measures in the Whole Sample

#### 3.2.2 Sex-Stratified Analysis

The associations of leptin with depressive episode (aOR=1.42; 95% CI 1.04-1.91), depressive symptoms (aOR=1.09; 95% CI 1.01-1.18) and negative symptoms (aOR=1.18; 95% CI 1.07-1.29) were stronger in women, as was the associations of adiponectin (aOR=1.07; 95% CI 1.01-1.13) and of insulin (aOR =1.08; 95% CI 1.01-1.14) with negative symptoms, although credible intervals overlapped with those for men (**Supplementary Tables 9&10**).

### 3.3 Associations of childhood immuno-metabolic factor scores with depression and psychosis risk in adulthood

#### 3.3.1 Whole Sample Analysis

The general immuno-metabolic factor, *G*, was associated with depression symptom score (OR=1.05; 95% CI 1.01-1.08) and psychotic experiences at age 24 (aOR=1.20; 95% CI 1.01-1.42, **Figure 3, Table 2**). The adiposity factor was associated with negative symptoms (aOR=1.07; 95% CI 1.02-1.12).

**Figure 3:**
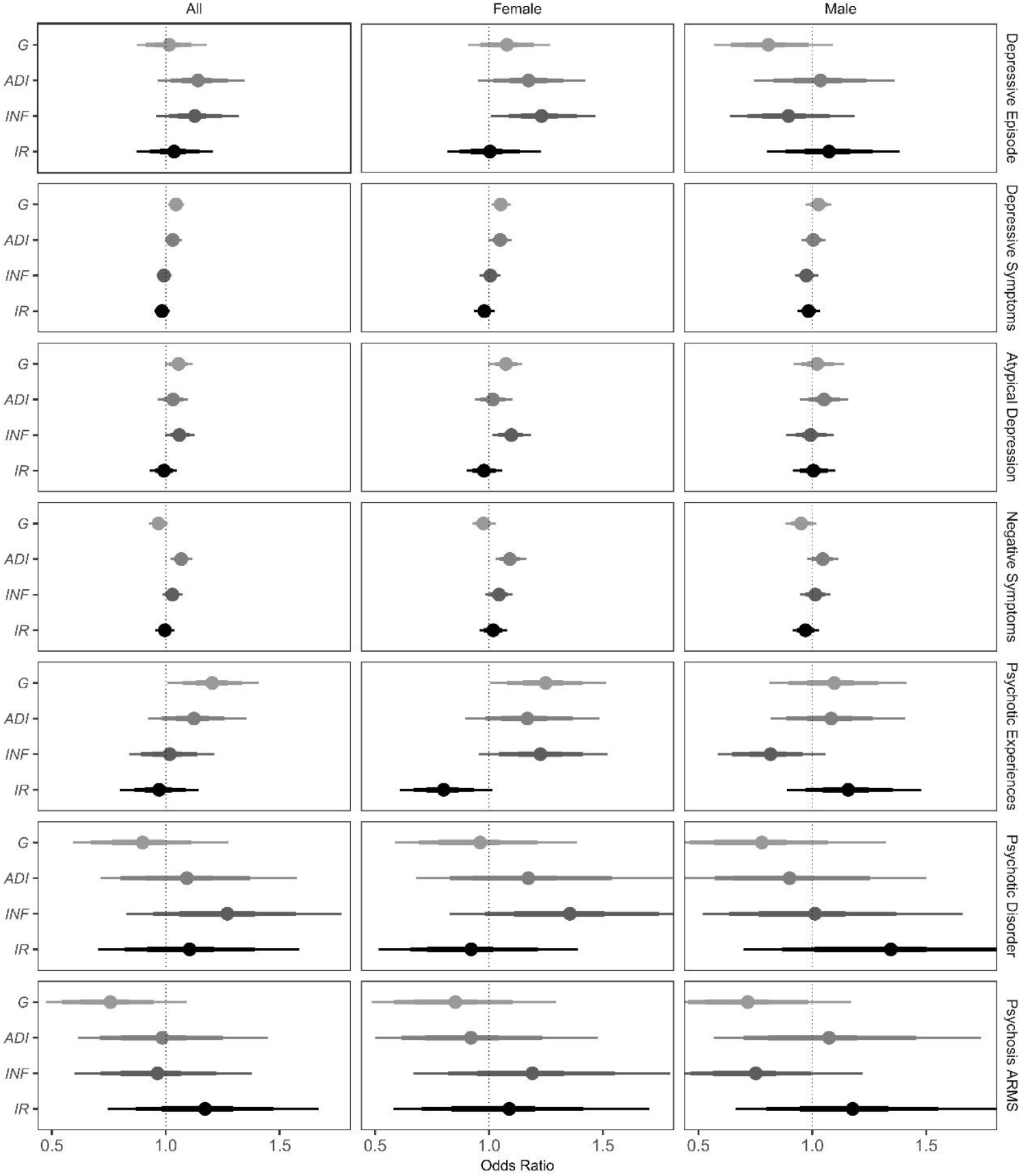
Associations of Childhood Immuno-Metabolic Factors with Adult Depression and Psychosis Measures in the Whole Sample. Plots of estimated odds ratios for the effect of factor scores derived from a bifactor EFA model (general factor (*G*) and three subfactors:(1) adiposity (*ADI*), (2) inflammation (*INF*) and (3) insulin resistance (*IR*) on psychiatric outcomes at age 24. Data are adjusted (for maternal social class and sex in the all-participant model) and grouped by model: either all participants, only females or only males. Thin bars represent the 95% credible interval, thicker bars the 66% credible interval, and dots the median of the posterior distribution for that parameter.

**Table 2:** Associations of Childhood Immuno-Metabolic Factors with Adult Depression and Psychosis Measures in the Whole Sample

| Outcome | Exposure<br>(Immuno-metabolic Factors) | Estimated OR<br>(95% CI) | pd | ROPE<br>(%) |
| --- | --- | --- | --- | --- |
| Depressive Episode | General factor | 1.02 (0.87, 1.18) | 0.843 | 79.33 |
|  | Adiposity factor | 1.14 (0.97, 1.35) | 0.121 | 34.71 |
|  | Inflammation factor | 1.13 (0.97, 1.33) | 0.141 | 40.11 |
|  | Insulin resistance factor | 1.04 (0.88, 1.22) | 0.670 | 73.37 |
| Depressive Symptoms<br>Score | General factor | 1.05 (1.01, 1.08) | 0.008 | 99.97 |
|  | Adiposity factor | 1.03 (0.99, 1.07) | 0.107 | 99.98 |
|  | Inflammation factor | 0.99 (0.96, 1.03) | 0.626 | 100.00 |
|  | Insulin resistance factor | 0.98 (0.95, 1.02) | 0.328 | 100.00 |
| Atypical Depressive<br>Symptom Score | General factor | 1.06 (1.00, 1.12) | 0.063 | 93.41 |
|  | Adiposity factor | 1.03 (0.97, 1.10) | 0.339 | 98.12 |
|  | Inflammation factor | 1.06 (1.00, 1.13) | 0.069 | 91.38 |
|  | Insulin resistance factor | 0.99 (0.93, 1.05) | 0.796 | 99.83 |
| Negative Symptoms<br>Score | General factor | 0.97 (0.93, 1.01) | 0.130 | 99.92 |
|  | Adiposity factor | 1.07 (1.02, 1.12) | 0.005 | 92.45 |
|  | Inflammation factor | 1.03 (0.99, 1.07) | 0.191 | 99.98 |
|  | Insulin resistance factor | 1.00 (0.95, 1.04) | 0.861 | 100.00 |
| Psychotic Experiences | General factor | 1.20 (1.01, 1.42) | 0.029 | 15.84 |
|  | Adiposity factor | 1.12 (0.93, 1.36) | 0.226 | 41.93 |
|  | Inflammation factor | 1.02 (0.85, 1.22) | 0.850 | 69.88 |
|  | Insulin resistance factor | 0.97 (0.81, 1.16) | 0.754 | 70.36 |
| Psychotic Disorder | General factor | 0.90 (0.62, 1.32) | 0.570 | 34.57 |
|  | Adiposity factor | 1.09 (0.72, 1.59) | 0.673 | 34.12 |
|  | Inflammation factor | 1.27 (0.86, 1.82) | 0.234 | 19.64 |

|  | Insulin resistance factor | 1.10 (0.75, 1.65) | 0.621 | 33.89 |
| --- | --- | --- | --- | --- |
| Psychosis ARMS | General factor | 0.76 (0.50, 1.13) | 0.171 | 16.25 |
|  | Adiposity factor | 0.98 (0.63, 1.47) | 0.942 | 35.86 |
|  | Inflammation factor | 0.96 (0.64, 1.44) | 0.854 | 36.54 |
|  | Insulin resistance factor | 1.17 (0.79, 1.73) | 0.427 | 28.33 |

#### 3.3.2 Sex-Stratified Analysis

There was stronger evidence of association between *G* and psychopathology in women than men. In women, *G* was associated with depression symptom score (aOR=1.05; 95% CI 1.01-1.09), atypical depression (aOR=1.07; 95% CI 1.00-1.15), and psychotic experiences (aOR=1.25; 95% CI 1.01-1.52, **Supplementary Tables 11&12**) although credible intervals overlapped with those for men.

The inflammatory factor was associated with depressive episodes (aOR=1.23; 95% CI 1.01-1.47) and atypical depressive symptoms in women (aOR=1.10; 95% CI 1.02-1.19). The adiposity factor was associated with negative symptoms in women (aOR=1.09; 95% CI 1.03-1.16). The insulin resistance factor was not associated with any psychiatric outcomes at age 24. However, point estimates for the association between the insulin resistance factor and psychosis-related outcomes (PEs, ARMS, psychotic disorder) were considerably larger in men compared with women, although credible intervals were relatively wide (**Figure 3**).

### 3.4 Missing Data

Out of total 15,645 participants enrolled in ALSPAC at inception, 3,875 contributed clinic and covariate data up to age 9, and up to 3,966 participants contributed psychiatric data at age 24 (**Supplementary Table 13**). A maximum of 1,611 participants contributed both exposure and outcome data (∼10% of the total cohort, ∼40% of all participants with outcome data). Therefore, there was substantial missingness in our dataset.

In a sample of participants who contributed all demographic, age 9 immuno-metabolic exposures and covariate data (n = 3,863), male sex (aOR=2.32; 95% CI 2.02-2.68), lower maternal education (aOR=0.39; 95% CI 0.29-0.50), lower maternal social class (aOR=0.62; 95% CI 0.40-0.95), and higher adiponectin levels at age 9 (aOR=1.08; 95% CI 1.01-1.16) were associated with missing psychiatric outcome data at age 24 (**Supplementary Table 14)**.

## 4. Discussion

### 4.1 Main Findings

Using data from a prospective birth cohort, we report that childhood leptin and adiponectin are associated with largely affective symptoms (depression and psychosis negative symptoms) in adulthood. Evidence for these associations remained after controlling for covariates including BMI and inflammation and was stronger in women. Exploratory Factor Analysis of metabolic and inflammatory biomarkers revealed an underlying structure of covariance, where a general immuno-metabolic factor was associated with both depressive symptoms and psychotic experiences. We also identified three specific factors. Leptin and BMI loaded onto the adiposity factor, which was associated with negative symptoms. The inflammatory factor was associated with depressive episode and atypical depressive symptoms in women. We found no associations between the insulin resistance factor and psychiatric outcomes, although the point estimates for psychosis-related outcomes were considerably larger in men than women.

### 4.2 Comparison with Previous Studies

The association between leptin and negative symptoms is in keeping with a meta-analysis of patients with schizophrenia (Stubbs et al., 2016), although this was not observed in smaller studies of first episode of psychosis (FEP) patients (Chouinard et al., 2019; Lis et al., 2020). However, in FEP cases Lis et al., (2020) observed a correlation between leptin and cognitive dysfunction, a domain of psychopathology somewhat linked to negative symptoms. Recently, a Mendelian randomisation (MR) study did not find an association between leptin and schizophrenia (Perry et al., 2021a), bringing into question whether leptin plays a causal role in the illness. It is worth noting that this MR analysis relied on GWAS of schizophrenia, and not that of negative symptoms specifically. Therefore, replication of our findings in other longitudinal cohorts, as well as evidence triangulation using complementary methods like MR based on GWAS of negative symptoms are required to fully understand the relationship between leptin and negative symptoms.

Lack of association between childhood random insulin levels, or the insulin resistance factor, and positive psychotic symptoms at age 24 is consistent with a previous study from the ALSPAC cohort testing association between fasting insulin and psychosis-spectrum outcomes at age 18 (Perry et al., 2019). However, using longitudinal repeat measure data, Perry et al., (2021b) recently reported that persistently raised fasting insulin levels through childhood and adolescence is associated with psychosis risk at age 24. Together, these findings suggest that persistently high leptin during development is likely relevant for adult psychosis, rather than one-off measures which do not consider variation in leptin levels over time. Nevertheless, similar to Perry et al., (2021b), we observed stronger association between the insulin resistance factor and psychosis-related outcomes in men compared with women, suggesting that early-life insulin resistance may contribute to psychosis risk in men particularly.

### 4.3 Potential mechanism of associations of immuno-metabolic markers and depressive and psychotic symptoms

We observed that adiposity and inflammation were largely associated with negative and atypical depressive symptoms. There is a rich literature suggesting a role of low-grade systemic inflammation in depression (Khandaker et al., 2017; Milaneschi et al., 2021, 2020). Our findings are consistent with a previous genetic study showing that cases of atypical depression (defined by increased appetite and/or weight) have higher genetic predisposition for obesity and inflammation, particularly genetic risk variants for BMI, leptin, and CRP (Milaneschi et al., 2017). Similarly, recent MR studies from the UK Biobank and Dutch NESDA cohorts have reported associations of higher BMI and IL-6 with specific depressive symptoms such as fatigue and appetite change (Kappelmann et al., 2021; Milaneschi et al., 2021). The mechanism of the association between inflammation and depression, particularly atypical depressive symptoms (Lamers et al., 2020), has been hypothesised to involve the activation of sickness-related behaviours by inflammatory cytokines, which directly affect neuroendocrine systems involved in regulating mood and homeostatic functions such as sleep and appetite (Milaneschi et al., 2020). Further work is needed to understand how leptin, and adiposity in general, contribute to negative symptoms.

Negative symptoms are common in schizophrenia, with prevalence exceeding 50% (Bobes et al., 2010), and are a major contributor to morbidity (Fervaha et al., 2014; Iasevoli et al., 2018). Furthermore, there is only a limited evidence base for effective treatments for these symptoms (Galderisi et al., 2021). Previous work suggests two latent factors underlying negative symptoms, namely reduced expression and amotivation (Bucci and Galderisi, 2017). Studies in schizophrenia suggest an association between amotivational negative symptoms and altered reward processing, which involves brain regions associated with the dopamine system including the ventral striatum and medial prefrontal cortex (Strauss et al., 2016; Vanes et al., 2018; Wang et al., 2015; Wolf et al., 2014). Leptin has been demonstrated to both indirectly and directly affect the activity of midbrain dopamine neurons in animal models (Hommel et al., 2006; Leinninger et al., 2009), as has adiponectin (Sun et al., 2019). Furthermore, a relationship between inflammation, brain reward systems and negative symptoms has been proposed (Goldsmith and Rapaport, 2020), which leptin could also modulate, given evidence of its interactions with immune system (Abella et al., 2017; Pulito-Cueto et al., 2020). Therefore, adipokines could have a direct influence on immune and neural networks involved in the pathogenesis of negative symptoms.

### 4.4 Strengths and Limitations

We used prospective data with exposures measured approximately 15 years prior to that of outcomes. We used a Bayesian approach to our statistical analysis, which is relatively novel and allowed the incorporation of prior information, and the generation of point estimates and credible intervals which have a more intuitive and robust interpretation than frequentist confidence intervals and p-values (Wagenmakers et al., 2018).

Limitations include missing data and pattern of missingness. Participants were not missing completely at random. Analysis of missingness in exposure data revealed that social class, and education influenced participation in the study. The complete case data came from a group from higher social class and higher educated families. One of the exposures, adiponectin, was associated with missingness, which might have introduced bias. Furthermore, >97% of the ALSPAC cohort is of white ethnicity, and so representativeness is an issue, and further studies in more diverse populations are required.

We used an exploratory factor analysis approach to derive immuno-metabolic factors. In future, confirmation of these findings in an independent dataset using confirmatory factor analysis is required. Bifactor models have a tendency to overfitting (Bonifay and Cai, 2017) and we were limited to nine immuno-metabolic biomarkers available from the age 9 ALSPAC cohort clinic assessment. In future, inclusion of a wider range of biomarkers may help to improve latent factor estimation. Nevertheless, without pre-specifying the biomarkers that might contribute to each latent factor, we identified three specific factors (adiposity, inflammation, and insulin resistance) composed of individual biomarkers with established, intuitive links with each factor, giving our findings face validity.

Methods for calculating factor scores are imprecise, and scores are inherently indeterminate (Grice, 2001). Therefore, the use of extracted factor scores as exposures in separate models may have introduced error. However, the approach of combining factor scores with other datasets for further analysis, including by Bayesian approaches, has been previously applied (Dam et al., 2017; Gagne et al., 2020), and can contribute exploratory information to inform future studies. A future approach that combines (potentially longitudinally measured) biomarkers with latent factors and psychiatric outcomes in a single (Bayesian) structural equation model might inform a deeper understanding of the relationship between individual biomarkers, useful intermediate concepts (latent factors), and individual symptoms or psychopathological constructs.

### 4.5 Conclusions

Using a prospective cohort study, we report that childhood levels of the metabolic hormones leptin and adiponectin are associated with depressive and negative symptoms in early adulthood. These associations are stronger in women than men. Somewhat distinct associations were seen for immuno-metabolic factor scores, which were derived from nine immuno-metabolic measures. While a general immuno-metabolic factor was associated with both depressive and psychotic symptoms, inflammation and adiposity were particularly associated with affective (depressive, atypical depressive, and negative) symptoms. Together, these findings are consistent with the idea that childhood immuno-metabolic alterations may contribute to risk of depressive and psychotic disorders in adulthood.

## Acknowledgements

The authors would like to thank Nils Kappelmann for valuable discussions about statistical analysis and atypical depression. We are extremely grateful to all the families who took part in this study, the midwives for their help in recruiting them, and the whole ALSPAC team, which includes interviewers, computer and laboratory technicians, clerical workers, research scientists, volunteers, managers, receptionists and nurses. The views expressed are those of the authors and not necessarily those of the NIHR or the Department of Health and Social Care.

## CRediT author statement

Nicholas Donnelly: Conceptualisation; Software; Data Curation; Formal Analysis; Visualization; Writing – Original Draft

Benjamin Perry: Data Curation; Writing – Review & Editing Hannah Jones: Writing – Reviewing & Editing

Golam Khandaker: Conceptualization; Supervision; Writing – Reviewing & Editing; Funding acquisition

