## Supplementary Materials for "Childhood Immuno-metabolic Markers and Risk of Depression and Psychosis in Adulthood: A Prospective Birth Cohort Study"

^2^ Avon and Wiltshire NHS Mental Health Partnership Trust

^3^ Department of Psychiatry, University of Cambridge

^4^ Cambridgeshire and Peterborough NHS Trust

^5^ NIHR Bristol Biomedical Research Centre, University of Bristol, Bristol, UK

^6^ MRC Integrative Epidemiology Unit, University of Bristol

### Supplementary Methods

#### Exploratory Factor Analysis

Suitability of our dataset for exploratory factor analysis (EFA) was tested using Bartlett’s test of sphericity (Χ^2^(36) = 7265.69, p < 0.001) and the Kaiser-Meyer-Olkin criterion (KMO, value = 0.656), which showed our data was suitable for EFA. Bartlett’s test compares an observed correlation matrix to an identity matrix (where there is no correlation between any variable); the KMO criterion estimates the degree to which each variable is predicted by other variables in a dataset. A value for the KMO > 0.5 is considered adequate for factor analysis (Kaiser and Rice, 1974). These measures indicate that our dataset was suitable for factor analysis.

Exploratory Factor Analysis was done using the *EFAtools* package (Steiner and Grieder, 2020) in R. The number of factors was selected after inspecting the results of an initial principal components analysis, where three components had eigenvalues > 1 (**Supplementary Figure 2A**). However, one eigenvalue was particularly large, suggesting the possible presence of a single underlying factor, which would also be in keeping with the established theoretical suggestion of a common “metabolic” factor. Therefore, we fit one factor, three factor, and a bifactor model with a general factor and 3 subfactors, using *EFA* with maximum likelihood estimation*.* For the bifactor model we used the “*bifactorQ*” rotation (Jennrich and Bentler, 2011; Mansolf and Reise, 2016). Similar results were obtained using the Schmid-Leiman transformation, an alternative method for exploratory bifactor EFA, using the *omega* function in the *psych* package (Revelle, 2013).

Models fit with 1 factor, 3 factors and the bifactor structure were compared with the Akaike Information Criteria (AIC), Bayesian Information Criteria (BIC), root mean square error of approximation (RMSEA), comparative fit index (CFI) and common part accounted for (CAF) measures. Compared to a one- or 3-factor model, a bifactor model with one general factor and three subfactors was a better fit to our data (**Supplementary** **Table 2**). Although concerns have been raised about the propensity for the bifactor model to overfit to data, (Bonifay and Cai, 2017), the factor structure produced had strong biological face validity for an exploratory analysis; we therefore took this structure forward for further analysis.

As a measure of the reliability of our factor estimates, we calculated McDonald’s omega total (ω_T_) statistic (McDonald, 1970; Zinbarg *et al.*, 2005; McNeish, 2018) for the bifactor model using the *OMEGA* function: general factor ω_T_ = 0.75, adiposity factor ω_T_ = 0.88, inflammatory factor ω_T_ = 0.65, insulin resistance factor ω_T_ = 0.78. Values < 0.6 are considered poor for measures of internal consistency (McNeish, 2018), we therefore considered our factor measures to be satisfactory.

Factor scores were extracted for all participants by the “*tenBerge*” method as this method is suited to EFA with oblique rotation (ten Berge *et al.*, 1999; Grice, 2001), **see Supplementary Table 3 and Supplementary Figure 2B for factor loadings**.

#### Bayesian Regression Model Fitting

We analysed data using R version 4.1.0 (R Development Core Team, 2017), fitting Bayesian regression models using the *brms* (Bürkner, 2017) and *rstanarm* packages, which fit models with Hamiltonian Monte Carlo using the *Stan* Language (Carpenter *et al.*, 2017).

We chose to fit Bayesian regression models as this approach has several advantages over frequentist methods including the ability to incorporate prior information into model estimation, which improves the accuracy of estimation, and more intuitive interpretations of Bayesian model estimates compared to their frequentist equivalents (Kruschke and Liddell, 2018; McElreath, 2018; Vandekerckhove *et al.*, 2018; Wagenmakers *et al.*, 2018). For example, The Bayesian *credible interval* is analogous to the frequentist *confidence interval* but describes the actual distribution of observed model posterior values, and therefore is more intuitive, as opposite to the confidence interval, which is inferred from theoretical characteristics of a particular distribution. Furthermore, the Bayesian approach provides alternatives to the use of p-values for null hypothesis significance testing and a focus on the idea of statistical significance, which has been much criticised e.g. Wagenmakers *et al.*, (2018).

Models with continuous outcomes (i.e. models examining the correlates of adiponectin, insulin or leptin) were fit with a Gaussian distribution and identity link function; binary outcomes were fit using a Bernoulli distribution and a log link function (Depressive Episode, Psychotic Experiences, Psychotic Disorder and Psychosis ARMS outcome models); count data was fit with Negative Binomial models (to account for overdispersion) and a log link function (Depressive Symptoms outcome models); and aggregated binomial data were modelled using a binomial distribution and logit link function (Atypical Depression and Negative Symptoms outcome models).

We used regularising priors for all Bayesian models as described by McElreath (2018); for exposure-outcome analysis models, priors for model beta coefficients were a normal distribution of mean = 0, and standard deviation (SD) = 0.5, prior for model intercepts were a normal distribution of mean = 0, and SD = 1.5 and priors for model error components were exponential distributions with rate = 1.

For all models estimates we calculated the 95% credible interval (CI), using the median and the highest density interval (HDI) method. The HDI is the interval where all points have a higher probability density than those outside the interval.

All models were fit with 16,000 posterior samples (4,000 warmup), and measures of model convergence were manually inspected. We calculated the Gelman-Rubin scale reduction statistic, *Rhat*, the ratio of variance within samples from a chain compared to the variance of all samples, values < 1.1 are considered acceptable; and the Effective Sample Size, a measure of the degree of autocorrelation within chains, values > 1000 are considered acceptable. All model values were within conventionally accepted limits.

As the number of participants providing data for different outcomes varied, we fit each model to the largest possible dataset of complete cases for all covariates, exposures and outcomes (**see Supplementary Tables 2-4 for full details**).

We fit unadjusted models with the outcome predicted by our exposure variables of interest (leptin, adiponectin and insulin). Next, we fit adjusted models with BMI at age 9, log_10_ IL-6, sex at birth and maternal socioeconomic class included as covariates. For models fit to sex-stratified data, we did not include sex at birth as a covariate. For models using factor scores as exposures, we included factor scores plus sex at birth and maternal socio-economic class as covariates.

We present as summary statistics the *probability of direction (pd)*, the proportion of a posterior distribution with the same sign as the posterior median (which can then be converted into a p-value analogue), which can be used as evidence for the existence of an effect; and the Region of Practical Equivalence (ROPE), the proportion of the posterior falling within a set region around the null, which can be used in interpreting is an effect is meaningfully different to the null, here defined as an odds ratio in the range [0.9 – 1.1] (Kruschke and Liddell, 2018; Makowski *et al.*, 2019a, 2019b).

### Supplementary Figures

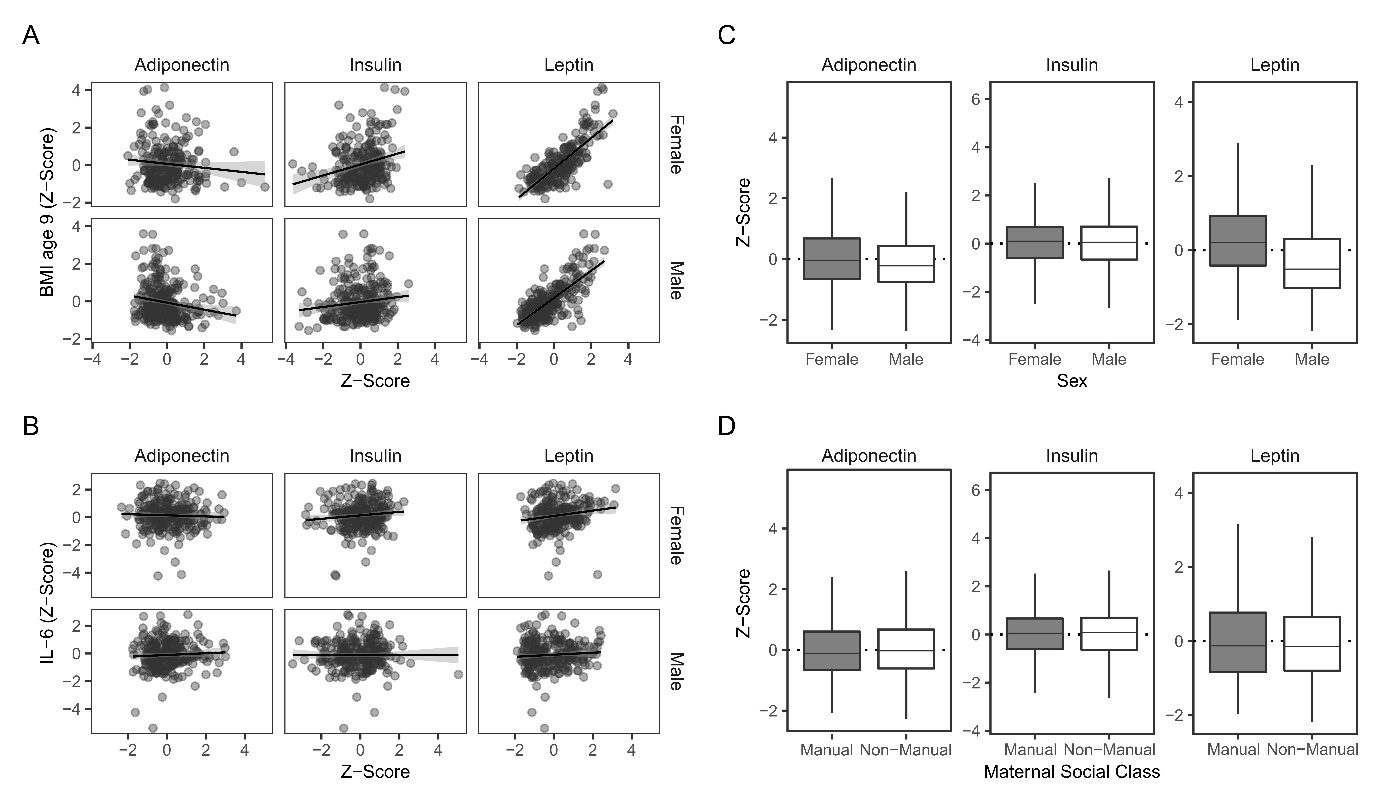

#### Supplementary Figure 1: Correlations of exposure variables and covariates

A. Scatter plot of (z-scored) adiponectin, random insulin or leptin against BMI, for female and male participants, with least squares regression line and its 95% confidence interval. Note that for interpretability of the plot, a randomly selected sample of 500 participants are plotted
B. Scatter plot of metabolic exposure variables by IL-6, as (A)
C. Box and whisker plots of metabolic exposure variables by sex. Boxes represent the median and IQR, whiskers represent 1.5 x the IQR
D. Box and whisker plots of metabolic exposure variables by maternal social class, as (C)

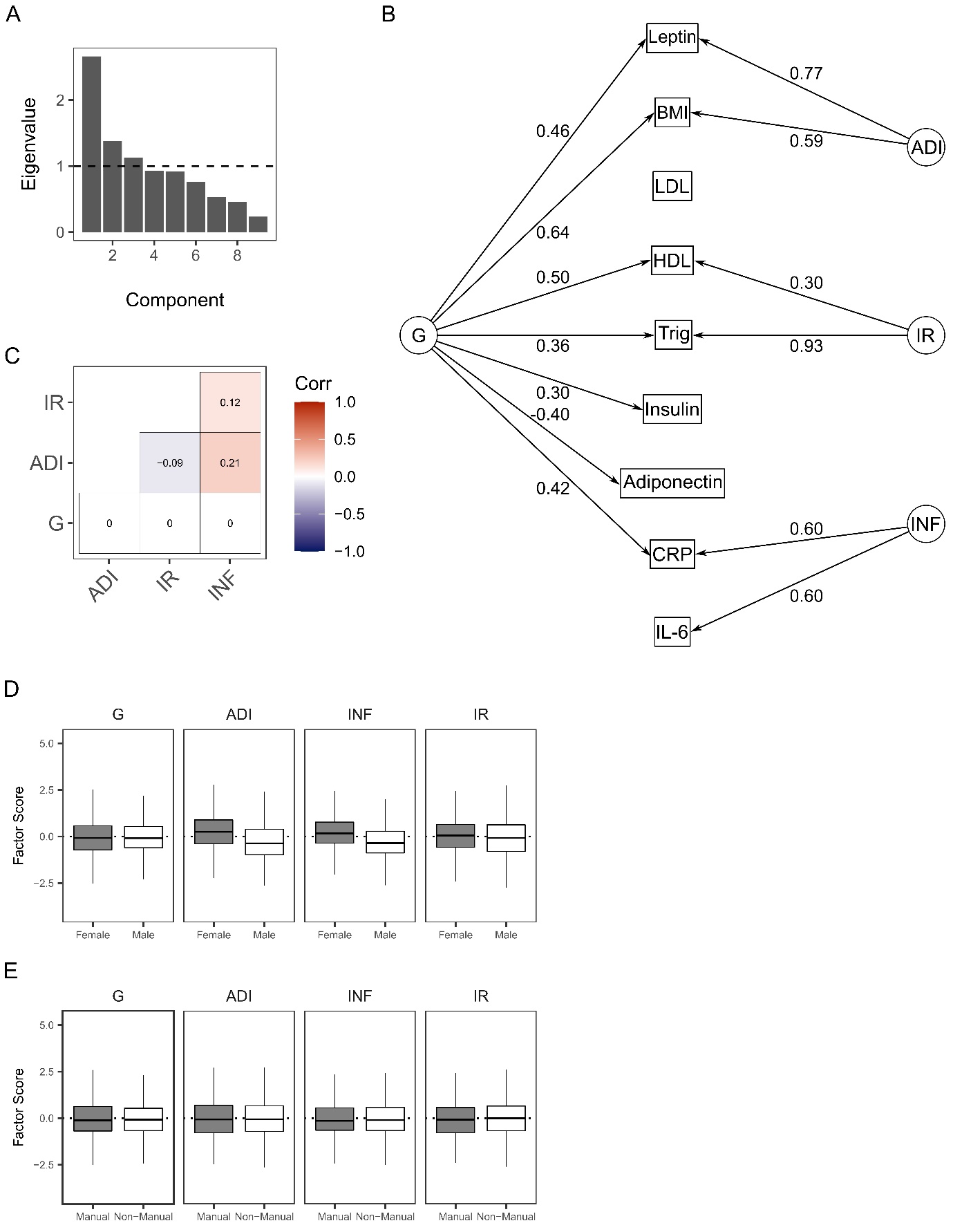

#### Supplementary Figure 2: Exploratory Factor Analysis characteristics

A. Bar plot of Eigenvalues extracted from principal components analysis of nine immuno-metabolic markers. Horizontal line indicates an eigenvalue of 1; components with an eigenvalue > 1 have greater power to explain variance in the dataset than individual variables, therefore we selected a number of factors equal to the number of components with eigenvalues > 1, in this case 3.
B. Path diagram of factor loadings (numbers) for the general factor (*G*) and three subfactors (adiposity factor, *ADI*; inflammatory factor, *INF* and insulin resistance factor, *IR*) on all nine biomarkers: leptin; BMI (Body Mass Index); LDL (low density lipoprotein); HDL (high density lipoprotein); Trig (triglycerides); insulin; adiponectin; CRP (C-reactive protein) and IL-6 (interleukin 6). Only paths with absolute loading values ≥ 0.30 are plotted.
C. Plot of correlations between the general factor (*G*) and three subfactors (adiposity factor, *ADI*; inflammatory factor, *INF* and insulin resistance Factor, *IR*). Colours indicate strength of correlation. Numerical values are correlation coefficients. Note a positive correlation between the inflammatory factor and both the adiposity and inflammatory factors, a negative correlation between the insulin resistance and adiposity factors, and minimal correlation between the general factor and any subfactor.
D. Box and whisker plots of factor scores by sex. Boxes represent the median and IQR, whiskers represent 1.5 x the IQR. Horizontal line indicates a score of zero.
E. Box and whisker plots of factor scores by maternal social class, as (D)

### Supplementary Tables

#### Supplementary Table 1: Immuno-metabolic marker characteristics at age 9

|  |  | **Sex** |  |
| --- | --- | --- | --- |
| **Measure** | **Overall (n = 3,875)** | **Female (n = 1,886)** | **Male (n = 1,989)** |
| **Adiponectin (µg/l)** | 1.24 (0.92, 1.62) | 1.29 (0.95, 1.69) | 1.19 (0.90, 1.55) |
| **Leptin (ng/ml)** | 5.00 (3.00, 10.00) | 7.00 (4.00, 12.00) | 4.00 (3.00, 8.00) |
| **Insulin (mU/l)** | 8.00 (4.00, 15.00) | 8.00 (4.00, 15.00) | 8.00 (4.00, 15.00) |
| **CRP (mg/l)** | 0.21 (0.11, 0.52) | 0.28 (0.14, 0.68) | 0.16 (0.10, 0.38) |
| **IL-6 (pg/ml)** | 0.78 (0.48, 1.37) | 0.89 (0.55, 1.50) | 0.70 (0.44, 1.20) |
| **Triglycerides (mmol/l)** | 1.01 (0.77, 1.38) | 1.04 (0.79, 1.39) | 0.99 (0.74, 1.38) |
| **HDL (mmol/l)** | 1.38 (1.19, 1.60) | 1.35 (1.16, 1.57) | 1.41 (1.22, 1.63) |
| **LDL (mmol/l)** | 2.31 (1.96, 2.69) | 2.41 (2.05, 2.78) | 2.22 (1.87, 2.60) |
| **BMI (kg/m²)** | 16.91 (15.63, 18.78) | 17.16 (15.69, 19.02) | 16.76 (15.59, 18.51) |
| Values are median (IQR) |  |  |  |

#### Supplementary Table 2: EFA Model Comparison

| **Model** | **RMSEA (90% CI)** | **AIC** | **BIC** | **CFI** | **CAF** | **ChiSq** | **DF** | **P-Value** |
| --- | --- | --- | --- | --- | --- | --- | --- | --- |
| **1 Factor** | 0.14 (0.14, 0.15) | 2051.44 | 1882.36 | 0.74 | 0.47 | 2105.40 | 27 | < 0.001 |
| **3 Factor** | 0.07 (0.07, 0.08) | 239.89 | 164.74 | 0.97 | 0.49 | 263.89 | 12 | < 0.001 |
| **Bifactor** | 0.04 (0.03, 0.06) | 38.31 | 0.73 | 0.99 | 0.50 | 50.31 | 6 | < 0.001 |

#### Supplementary Table 3: EFA Bifactor Model Variable Loadings

| **Measure** | **General** | **Adiposity** | **Inflammatory** | **Insulin**  **Resistance** | **Communality (h2)** |
| --- | --- | --- | --- | --- | --- |
| **Adiponectin** | **-0.40** | 0.15 | 0.15 | 0.05 | 0.22 |
| **Leptin** | **0.46** | **0.77** | 0.06 | 0.03 | 0.84 |
| **Insulin** | **0.30** | 0.06 | -0.08 | 0.13 | 0.12 |
| **CRP** | **0.42** | 0.10 | **0.60** | -0.05 | 0.58 |
| **IL-6** | 0.27 | -0.04 | **0.60** | 0.02 | 0.43 |
| **Triglycerides** | **0.36** | 0.01 | -0.03 | **0.93** | 1.00 |
| **HDL** | **0.50** | -0.06 | 0.12 | **0.30** | 0.35 |
| **LDL** | -0.04 | 0.22 | -0.01 | 0.00 | 0.05 |
| **BMI** | **0.64** | **0.59** | -0.02 | -0.07 | 0.75 |

#### Supplementary Table 4: Depression Outcomes

|  |  | **Depressive Episode** |  |
| --- | --- | --- | --- |
| **Variable** | **Overall**  **(n = 1,808)** | **Absent**  **(n = 1,639)** | **Present**  **(n = 169)** |
| Depressive Symptoms | 5.00 (2.00, 8.00) | 4.00 (2.0, 7.0) | 7.00 (4.0, 11.0) |
| Atypical Depression Score | 1.00 (0.00, 1.00) | 0.00 (0.00, 1.00) | 1.00 (1.00, 2.00) |
| Leptin (ng/ml) | 5.50 (3.40, 10.03) | 5.40 (3.35, 9.80) | 7.00 (4.10, 12.10) |
| Adiponectin (µg/l) | 12.37 (9.11, 16.22) | 12.34 (9.09, 16.26) | 12.58 (9.55, 15.67) |
| Insulin (mU/l) | 8.29 (4.08, 15.29) | 8.17 (4.04, 15.30) | 9.12 (4.37, 14.70) |
| IL-6 (pg/ml) | 0.78 (0.48, 1.42) | 0.76 (0.47, 1.40) | 0.96 (0.57, 1.77) |
| BMI age 9 (kg/m²) | 16.90 (15.61, 18.68) | 16.89 (15.59, 18.66) | 17.04 (15.72, 19.14) |
| Sex |  |  |  |
| Female | 1,077 (60%) | 953 (58%) | 124 (73%) |
| Male | 731 (40%) | 686 (42%) | 45 (27%) |
| Maternal Social Class |  |  |  |
| Manual | 228 (13%) | 199 (12%) | 29 (17%) |
| Non-Manual | 1,580 (87%) | 1,440 (88%) | 140 (83%) |
| Median (IQR); n (%) |  |  |  |

#### Supplementary Table 5: Psychosis Outcomes

|  |  | **Psychotic Experiences** | | **Psychotic Disorder** | | **Psychosis ARMS** |  |
| --- | --- | --- | --- | --- | --- | --- | --- |
| **Variable** | **Overall (n = 1,776)** | **Absent (n = 1,646)** | **Present (n = 130)** | **Absent (n = 1,756)** | **Present (n = 20)** | **Absent (n = 1,756)** | **Present (n = 20)** |
| Leptin (ng/ml) | 5.50 (3.40, 10.12) | 5.50 (3.40, 10.00) | 6.15 (3.62, 11.75) | 5.50 (3.40, 10.10) | 7.65 (3.58, 11.68) | 5.50 (3.40, 10.12) | 5.70 (3.27, 9.67) |
| Adiponectin (µg/l) | 12.37 (9.13, 16.17) | 12.38 (9.13, 16.24) | 12.08 (9.24, 15.42) | 12.37 (9.10, 16.19) | 12.35 (10.28, 15.76) | 12.37 (9.12, 16.16) | 13.36 (10.04, 16.55) |
| Insulin (mU/l) | 8.31 (4.06, 15.27) | 8.18 (3.98, 15.06) | 10.75 (4.97, 18.51) | 8.30 (4.05, 15.21) | 9.89 (4.73, 19.21) | 8.31 (4.07, 15.21) | 7.07 (2.39, 17.85) |
| IL-6 (pg/ml) | 0.79 (0.48, 1.42) | 0.78 (0.48, 1.40) | 0.91 (0.51, 1.69) | 0.78 (0.48, 1.42) | 1.13 (0.62, 2.52) | 0.78 (0.48, 1.43) | 0.89 (0.48, 1.20) |
| BMI age 9 (kg/m²) | 16.91 (15.60, 18.71) | 16.88 (15.55, 18.70) | 17.42 (16.04, 18.88) | 16.91 (15.60, 18.71) | 17.01 (16.07, 18.13) | 16.92 (15.61, 18.72) | 16.18 (15.52, 17.32) |
| Sex |  |  |  |  |  |  |  |
| Female | 1,058 (60%) | 984 (60%) | 74 (57%) | 1,044 (59%) | 14 (70%) | 1,047 (60%) | 11 (55%) |
| Male | 718 (40%) | 662 (40%) | 56 (43%) | 712 (41%) | 6 (30%) | 709 (40%) | 9 (45%) |
| Maternal Social Class |  |  |  |  |  |  |  |
| Manual | 226 (13%) | 207 (13%) | 19 (15%) | 222 (13%) | <5 (20%) | 221 (13%) | 5 (25%) |
| Non-Manual | 1,550 (87%) | 1,439 (87%) | 111 (85%) | 1,534 (87%) | 16 (80%) | 1,535 (87%) | 15 (75%) |
| Median (IQR); n (%) |  |  |  |  |  |  |  |

#### Supplementary Table 6: Negative Symptoms

|  |  | **Negative Symptoms (n)** | | | | | | | | | | |
| --- | --- | --- | --- | --- | --- | --- | --- | --- | --- | --- | --- | --- |
| **Variable** | **Overall**  **(n = 1,753)** | **0**  **(n = 928)** | **1**  **(n = 287)** | **2**  **(n = 147)** | **3**  **(n = 111)** | **4**  **(n = 83)** | **5**  **(n = 62)** | **6**  **(n = 45)** | **7**  **(n = 30)** | **8**  **(n = 26)** | **9**  **(n = 23)** | **10**  **(n = 11)** |
| Leptin (ng/ml) | 5.50 (3.40, 10.10) | 5.60 (3.48, 10.10) | 5.40 (3.50, 9.70) | 4.80 (2.95, 9.05) | 6.00 (4.00, 11.20) | 5.10 (3.10, 9.65) | 5.45 (3.52, 11.00) | 5.10 (3.20, 7.70) | 4.55 (3.92, 10.78) | 4.80 (3.80, 10.40) | 7.90 (4.00, 14.90) | 11.40 (6.15, 16.70) |
| Adiponectin (µg/l) | 12.37 (9.10, 16.19) | 12.41 (9.22, 15.93) | 12.41 (9.18, 16.91) | 11.63 (8.62, 15.76) | 12.37 (9.42, 16.22) | 12.03 (8.28, 15.10) | 13.01 (8.83, 17.18) | 12.50 (8.59, 17.22) | 12.82 (9.70, 14.49) | 12.82 (9.58, 16.63) | 12.55 (9.37, 16.91) | 13.56 (12.29, 19.89) |
| Insulin (mU/l) | 8.29 (4.10, 15.32) | 8.41 (4.50, 15.30) | 7.52 (3.51, 15.85) | 7.72 (3.66, 13.73) | 7.73 (3.66, 14.96) | 11.07 (4.32, 16.51) | 9.65 (4.16, 17.32) | 6.89 (3.55, 15.31) | 9.32 (5.40, 15.90) | 4.46 (3.27, 11.40) | 11.37 (4.91, 15.44) | 7.43 (4.56, 12.94) |
| IL-6 (pg/ml) | 0.78 (0.48, 1.42) | 0.77 (0.47, 1.36) | 0.82 (0.51, 1.48) | 0.70 (0.42, 1.37) | 0.74 (0.51, 1.25) | 0.82 (0.44, 1.33) | 0.93 (0.50, 1.66) | 0.90 (0.56, 1.73) | 0.88 (0.52, 1.93) | 0.89 (0.53, 1.26) | 0.87 (0.60, 1.49) | 1.34 (0.46, 1.82) |
| BMI age 9 (kg/m²) | 16.91 (15.63, 18.71) | 17.03 (15.68, 18.75) | 16.78 (15.67, 18.63) | 16.70 (15.43, 18.24) | 17.17 (15.87, 18.92) | 16.73 (15.69, 18.38) | 16.41 (15.05, 18.21) | 16.68 (15.65, 18.09) | 16.58 (15.34, 17.67) | 16.60 (15.10, 18.92) | 17.96 (16.02, 20.87) | 18.71 (17.32, 20.17) |
| Sex |  |  |  |  |  |  |  |  |  |  |  |  |
| Female | 1,043 (59%) | 583 (63%) | 163 (57%) | 76 (52%) | 65 (59%) | 38 (46%) | 35 (56%) | 24 (53%) | 20 (67%) | 18 (69%) | 15 (65%) | 6 (55%) |
| Male | 710 (41%) | 345 (37%) | 124 (43%) | 71 (48%) | 46 (41%) | 45 (54%) | 27 (44%) | 21 (47%) | 10 (33%) | 8 (31%) | 8 (35%) | 5 (45%) |
| Maternal Social Class |  |  |  |  |  |  |  |  |  |  |  |  |
| Manual | 219 (12%) | 99 (11%) | 42 (15%) | 22 (15%) | 13 (12%) | 14 (17%) | 10 (16%) | 9 (20%) | <5 (6.7%) | <5 (7.7%) | <5 (17%) | <5 (18%) |
| Non-Manual | 1,534 (88%) | 829 (89%) | 245 (85%) | 125 (85%) | 98 (88%) | 69 (83%) | 52 (84%) | 36 (80%) | 28 (93%) | 24 (92%) | 19 (83%) | 9 (82%) |
| Median (IQR); n (%) |  |  |  |  |  |  |  |  |  |  |  |  |

#### Supplementary Table 7: Metabolic Exposure – Covariate Relationships

| **Outcome** | **Variable** |  | **Estimate (95% CI)** | **pd** |
| --- | --- | --- | --- | --- |
| Adiponectin | BMI |  | -0.14 (-0.16, -0.11) | < 0.001 |
|  | IL-6 |  | 0.04 (0.01, 0.06) | 0.007 |
|  | Maternal Social Class | Manual | *Reference* |  |
|  |  | Non-Manual | 0.04 (-0.03, 0.11) | 0.308 |
|  | Sex | Male | *Reference* |  |
|  |  | Female | 0.12 (0.07, 0.18) | < 0.001 |
| Insulin | BMI |  | 0.22 (0.19, 0.25) | < 0.001 |
|  | IL-6 |  | -0.01 (-0.04, 0.02) | 0.458 |
|  | Maternal Social Class | Manual | *Reference* |  |
|  |  | Non-Manual | 0.00 (-0.07, 0.09) | 0.939 |
|  | Sex | Male | *Reference* |  |
|  |  | Female | 0.00 (-0.06, 0.06) | 0.935 |
| Leptin | BMI |  | 0.71 (0.69, 0.73) | < 0.001 |
|  | IL-6 |  | 0.04 (0.02, 0.05) | < 0.001 |
|  | Maternal Social Class | Manual | *Reference* |  |
|  |  | Non-Manual | 0.02 (-0.03, 0.07) | 0.458 |
|  | Sex | Male | *Reference* |  |
|  |  | Female | 0.48 (0.44, 0.52) | < 0.001 |

#### Supplementary Table 8: Factor Score – Covariate Models

| **Factor** | **Variable** |  | **Estimate (95% CI)** | **pd** |
| --- | --- | --- | --- | --- |
| *G* | Sex | Male | *Reference* |  |
|  |  | Female | 0.00 (-0.06, 0.06) | 0.964 |
|  | Maternal Social Class | Manual | *Reference* |  |
|  |  | Non-Manual | -0.07 (-0.16, 0.01) | 0.095 |
| *ADI* | Sex | Male | *Reference* |  |
|  |  | Female | 0.53 (0.47, 0.59) | < 0.001 |
|  | Maternal Social Class | Manual | *Reference* |  |
|  |  | Non-Manual | 0.02 (-0.07, 0.10) | 0.699 |
| *INF* | Sex | Male | *Reference* |  |
|  |  | Female | 0.48 (0.42, 0.55) | < 0.001 |
|  | Maternal Social Class | Manual | *Reference* |  |
|  |  | Non-Manual | 0.03 (-0.05, 0.12) | 0.405 |
| *IR* | Sex | Male | *Reference* |  |
|  |  | Female | 0.14 (0.08, 0.20) | < 0.001 |
|  | Maternal Social Class | Manual | *Reference* |  |
|  |  | Non-Manual | 0.07 (-0.02, 0.16) | 0.121 |

#### Supplementary Table 9: Female Participants Exposure – Outcome Models

|  | ***Unadjusted*** | |  |  | ***Adjusted*** |  |  |
| --- | --- | --- | --- | --- | --- | --- | --- |
| **Outcome** | **Parameter** | **Estimated OR**  **(95% CI)** | **pd** | **ROPE (%)** | **Estimated OR**  **(95% CI)** | **pd** | **ROPE (%)** |
| Depressive Episode | Adiponectin | 1.00 (0.83, 1.20) | 0.976 | 70.40 | 0.99 (0.81, 1.20) | 0.898 | 67.9 |
|  | Insulin | 1.10 (0.91, 1.34) | 0.315 | 48.98 | 1.13 (0.93, 1.37) | 0.231 | 41.03 |
|  | Leptin | 1.26 (1.03, 1.55) | 0.028 | 10.40 | 1.42 (1.04, 1.91) | 0.029 | 5.40 |
| Depressive Symptoms | Adiponectin | 0.97 (0.93, 1.01) | 0.122 | 99.81 | 0.97 (0.92, 1.01) | 0.140 | 99.76 |
|  | Insulin | 0.99 (0.95, 1.03) | 0.698 | 99.99 | 0.99 (0.95, 1.04) | 0.800 | 100.00 |
|  | Leptin | 1.08 (1.03, 1.13) | 0.001 | 83.37 | 1.09 (1.01, 1.18) | 0.022 | 63.46 |
| Atypical Depression | Adiponectin | 0.99 (0.91, 1.06) | 0.718 | 98.48 | 0.99 (0.92, 1.07) | 0.878 | 98.86 |
|  | Insulin | 1.02 (0.94, 1.10) | 0.649 | 97.94 | 1.02 (0.94, 1.11) | 0.666 | 97.94 |
|  | Leptin | 1.07 (0.99, 1.16) | 0.113 | 79.78 | 1.01 (0.89, 1.15) | 0.837 | 86.47 |
| Negative Symptoms | Adiponectin | 1.08 (1.02, 1.14) | 0.008 | 79.90 | 1.07 (1.01, 1.13) | 0.024 | 87.06 |
|  | Insulin | 1.06 (1.01, 1.13) | 0.047 | 90.42 | 1.08 (1.01, 1.14) | 0.012 | 80.19 |
|  | Leptin | 1.08 (1.02, 1.15) | 0.019 | 78.42 | 1.18 (1.07, 1.29) | 0.001 | 9.23 |
| Psychotic Experiences | Adiponectin | 0.98 (0.77, 1.24) | 0.888 | 58.18 | 1.01 (0.79, 1.29) | 0.915 | 57.48 |
|  | Insulin | 1.18 (0.93, 1.51) | 0.177 | 27.77 | 1.17 (0.90, 1.49) | 0.215 | 31.01 |
|  | Leptin | 1.29 (1.00, 1.64) | 0.047 | 11.22 | 1.06 (0.73, 1.53) | 0.737 | 38.93 |
| Psychotic Disorder | Adiponectin | 0.96 (0.61, 1.51) | 0.860 | 33.37 | 0.93 (0.58, 1.49) | 0.763 | 31.38 |
|  | Insulin | 1.17 (0.74, 1.84) | 0.512 | 26.96 | 1.21 (0.74, 1.90) | 0.412 | 24.05 |
|  | Leptin | 1.18 (0.75, 1.91) | 0.482 | 26.17 | 1.37 (0.76, 2.47) | 0.295 | 15.63 |
| Psychosis ARMS | Adiponectin | 0.89 (0.53, 1.43) | 0.631 | 28.42 | 0.85 (0.51, 1.40) | 0.519 | 25.40 |
|  | Insulin | 1.12 (0.68, 1.88) | 0.654 | 27.90 | 1.16 (0.72, 1.95) | 0.540 | 26.32 |
|  | Leptin | 0.92 (0.55, 1.55) | 0.757 | 28.36 | 1.18 (0.63, 2.18) | 0.601 | 21.78 |

#### Supplementary Table 10: Male Participants Exposure – Outcome Model

|  | ***Unadjusted*** | |  |  | ***Adjusted*** |  |  |
| --- | --- | --- | --- | --- | --- | --- | --- |
| **Outcome** | **Parameter** | **Estimated OR**  **(95% CI)** | **pd** | **ROPE (%)** | **Estimated OR**  **(95% CI)** | **pd** | **ROPE (%)** |
| Depressive Episode | Adiponectin | 1.10 (0.81, 1.45) | 0.547 | 41.66 | 1.09 (0.81, 1.46) | 0.548 | 42.33 |
|  | Insulin | 0.89 (0.68, 1.17) | 0.410 | 39.83 | 0.90 (0.69, 1.19) | 0.449 | 40.75 |
|  | Leptin | 0.94 (0.69, 1.28) | 0.711 | 45.12 | 0.97 (0.65, 1.44) | 0.879 | 36.33 |
| Depressive Symptoms | Adiponectin | 1.01 (0.96, 1.07) | 0.678 | 99.95 | 1.02 (0.97, 1.08) | 0.474 | 99.86 |
|  | Insulin | 1.04 (0.99, 1.10) | 0.107 | 98.89 | 1.04 (0.98, 1.09) | 0.176 | 99.35 |
|  | Leptin | 1.00 (0.94, 1.05) | 0.897 | 99.95 | 0.95 (0.88, 1.03) | 0.208 | 87.37 |
| Atypical Depression | Adiponectin | 1.01 (0.91, 1.12) | 0.900 | 94.07 | 1.01 (0.91, 1.12) | 0.789 | 92.67 |
|  | Insulin | 0.94 (0.86, 1.04) | 0.227 | 79.94 | 0.93 (0.85, 1.02) | 0.153 | 72.09 |
|  | Leptin | 1.06 (0.95, 1.17) | 0.300 | 80.75 | 0.96 (0.83, 1.12) | 0.650 | 75.05 |
| Negative Symptoms | Adiponectin | 1.04 (0.97, 1.11) | 0.235 | 96.58 | 1.04 (0.97, 1.11) | 0.300 | 97.02 |
|  | Insulin | 0.97 (0.91, 1.03) | 0.280 | 98.61 | 0.97 (0.91, 1.03) | 0.353 | 98.76 |
|  | Leptin | 1.04 (0.97, 1.11) | 0.281 | 96.92 | 1.07 (0.98, 1.19) | 0.164 | 71.03 |
| Psychotic Experiences | Adiponectin | 0.99 (0.75, 1.30) | 0.960 | 52.64 | 1.02 (0.78, 1.34) | 0.873 | 52.09 |
|  | Insulin | 1.11 (0.86, 1.44) | 0.439 | 43.29 | 1.09 (0.84, 1.41) | 0.511 | 46 |
|  | Leptin | 1.03 (0.79, 1.36) | 0.857 | 53.25 | 0.90 (0.62, 1.32) | 0.616 | 34.68 |
| Psychotic Disorder | Adiponectin | 1.19 (0.66, 2.04) | 0.540 | 22.35 | 1.18 (0.66, 2.04) | 0.588 | 22.47 |
|  | Insulin | 0.94 (0.56, 1.62) | 0.812 | 27.32 | 0.94 (0.57, 1.61) | 0.818 | 29.04 |
|  | Leptin | 0.83 (0.44, 1.56) | 0.555 | 20.77 | 0.85 (0.44, 1.67) | 0.645 | 20.34 |
| Psychosis ARMS | Adiponectin | 1.13 (0.67, 1.90) | 0.643 | 26.22 | 1.13 (0.66, 1.89) | 0.639 | 25.14 |
|  | Insulin | 0.73 (0.45, 1.17) | 0.190 | 13.74 | 0.73 (0.46, 1.19) | 0.214 | 14.81 |
|  | Leptin | 0.99 (0.56, 1.69) | 0.968 | 28.05 | 1.06 (0.57, 1.94) | 0.851 | 24.21 |

#### Supplementary Table 11: Female Participant Factor-Outcome Models

| **Outcome** | **Exposure**  **(Immuno-metabolic Factors)** | **Estimated OR (95% CI)** | **pd** | **ROPE (%)** |
| --- | --- | --- | --- | --- |
| Depressive Episode | General factor | 1.08 (0.91, 1.27) | 0.367 | 59.14 |
|  | Adiposity factor | 1.17 (0.96, 1.43) | 0.120 | 27.35 |
|  | Inflammation factor | 1.23 (1.01, 1.47) | 0.031 | 12.94 |
|  | Insulin resistance factor | 1.00 (0.82, 1.23) | 0.969 | 66.73 |
| Depressive Symptoms | General factor | 1.05 (1.01, 1.09) | 0.014 | 99.33 |
|  | Adiposity factor | 1.05 (1.00, 1.10) | 0.051 | 98.16 |
|  | Inflammation factor | 1.01 (0.96, 1.05) | 0.788 | 100.00 |
|  | Insulin resistance factor | 0.98 (0.93, 1.02) | 0.368 | 99.91 |
| Atypical Depression | General factor | 1.07 (1.00, 1.15) | 0.045 | 79.25 |
|  | Adiposity factor | 1.02 (0.94, 1.10) | 0.675 | 97.54 |
|  | Inflammation factor | 1.10 (1.02, 1.19) | 0.014 | 56.08 |
|  | Insulin resistance factor | 0.98 (0.9, 1.06) | 0.594 | 96.85 |
| Negative Symptoms | General factor | 0.97 (0.93, 1.03) | 0.352 | 99.72 |
|  | Adiposity factor | 1.09 (1.03, 1.16) | 0.005 | 65.42 |
|  | Inflammation factor | 1.04 (0.98, 1.10) | 0.148 | 97.47 |
|  | Insulin resistance factor | 1.02 (0.96, 1.08) | 0.562 | 99.54 |
| Psychotic Experiences | General factor | 1.25 (1.01, 1.52) | 0.038 | 12.58 |
|  | Adiposity factor | 1.17 (0.91, 1.51) | 0.227 | 31.07 |
|  | Inflammation factor | 1.23 (0.98, 1.55) | 0.087 | 18.12 |
|  | Insulin resistance factor | 0.80 (0.62, 1.04) | 0.085 | 17.15 |
| Psychotic Disorder | General factor | 0.96 (0.64, 1.47) | 0.854 | 35.90 |
|  | Adiposity factor | 1.17 (0.74, 1.90) | 0.494 | 26.01 |
|  | Inflammation factor | 1.35 (0.86, 2.03) | 0.182 | 14.15 |
|  | Insulin resistance factor | 0.92 (0.56, 1.46) | 0.727 | 30.68 |
| Psychosis ARMS | General factor | 0.85 (0.52, 1.34) | 0.515 | 26.62 |
|  | Adiposity factor | 0.92 (0.55, 1.57) | 0.764 | 27.15 |
|  | Inflammation factor | 1.19 (0.74, 1.92) | 0.475 | 24.64 |
|  | Insulin resistance factor | 1.09 (0.66, 1.83) | 0.740 | 28.37 |

#### Supplementary Table 12: Male Participant Factor-Outcome Models

| **Outcome** | **Exposure**  **(Immuno-metabolic Factors)** | **Estimated OR (95% CI)** | **pd** | **ROPE (%)** |
| --- | --- | --- | --- | --- |
| Depressive Episode | General factor | 0.81 (0.58, 1.11) | 0.185 | 21.14 |
|  | Adiposity factor | 1.04 (0.77, 1.41) | 0.817 | 47.12 |
|  | Inflammation factor | 0.90 (0.66, 1.22) | 0.476 | 38.74 |
|  | Insulin resistance factor | 1.07 (0.81, 1.40) | 0.626 | 46.93 |
| Depressive Symptoms | General factor | 1.03 (0.97, 1.08) | 0.340 | 99.48 |
|  | Adiposity factor | 1.00 (0.95, 1.06) | 0.879 | 99.97 |
|  | Inflammation factor | 0.97 (0.92, 1.03) | 0.336 | 99.62 |
|  | Insulin resistance factor | 0.98 (0.94, 1.03) | 0.521 | 99.98 |
| Atypical Depression | General factor | 1.02 (0.92, 1.14) | 0.712 | 90.97 |
|  | Adiposity factor | 1.05 (0.95, 1.16) | 0.337 | 83.30 |
|  | Inflammation factor | 0.99 (0.89, 1.10) | 0.872 | 93.66 |
|  | Insulin resistance factor | 1.01 (0.92, 1.10) | 0.914 | 96.60 |
| Negative Symptoms | General factor | 0.95 (0.88, 1.02) | 0.164 | 90.93 |
|  | Adiposity factor | 1.05 (0.98, 1.12) | 0.187 | 94.79 |
|  | Inflammation factor | 1.01 (0.95, 1.08) | 0.696 | 99.45 |
|  | Insulin resistance factor | 0.97 (0.91, 1.03) | 0.328 | 98.59 |
| Psychotic Experiences | General factor | 1.10 (0.83, 1.43) | 0.512 | 42.99 |
|  | Adiposity factor | 1.08 (0.83, 1.43) | 0.562 | 46.47 |
|  | Inflammation factor | 0.82 (0.61, 1.09) | 0.168 | 22.78 |
|  | Insulin resistance factor | 1.16 (0.90, 1.49) | 0.262 | 33.74 |
| Psychotic Disorder | General factor | 0.78 (0.42, 1.40) | 0.405 | 19.03 |
|  | Adiposity factor | 0.90 (0.49, 1.60) | 0.722 | 25.2 |
|  | Inflammation factor | 1.01 (0.58, 1.79) | 0.963 | 27.58 |
|  | Insulin resistance factor | 1.34 (0.77, 2.33) | 0.290 | 16.23 |
| Psychosis ARMS | General factor | 0.72 (0.40, 1.27) | 0.245 | 14.45 |
|  | Adiposity factor | 1.07 (0.61, 1.83) | 0.795 | 26.77 |
|  | Inflammation factor | 0.75 (0.42, 1.27) | 0.298 | 17.25 |
|  | Insulin resistance factor | 1.18 (0.71, 1.93) | 0.514 | 24.73 |

#### Supplementary Table 13: Missing Data

| **Outcome** | **With Outcome**  **(n, % Cohort)** | **With Outcome and Exposures**  **(n, % Cohort)** | **Outcome and Exposures**  **(% of all with Exposures)** |
| --- | --- | --- | --- |
| Depressive Episode | 3966 (25.35) | 1611 (10.30) | 40.62 |
| Depression Symptom Score | 3966 (25.35) | 1611 (10.30) | 40.62 |
| Atypical Depression Score | 3966 (25.35) | 1611 (10.30) | 40.62 |
| Psychotic Experiences Now | 3889 (24.86) | 1583 (10.12) | 40.70 |
| Psychotic Disorder Now | 3889 (24.86) | 1583 (10.12) | 40.70 |
| Psychosis ARMs | 3889 (24.86) | 1583 (10.12) | 40.70 |
| Negative Symptom Score | 3823 (24.44) | 1565 (10.00) | 40.94 |

#### Supplementary Table 14: Missing Outcomes Model

|  | **Variable** | **Estimate (95% CI)** | **pd** | **ROPE (%)** |
| --- | --- | --- | --- | --- |
| **Ethnicity** | White | *Reference* |  |  |
|  | Non-White | 0.97 (0.62, 1.58) | 0.896 | 32.55 |
| **Sex** | Female | *Reference* |  |  |
|  | Male | 2.32 (2.02, 2.68) | < 0.001 | 0.00 |
| **Maternal Education** | CSE/none | *Reference* |  |  |
|  | Vocational | 0.96 (0.70, 1.30) | 0.803 | 45.93 |
|  | O - Level | 0.61 (0.49, 0.77) | < 0.001 | 0.02 |
|  | A - Level | 0.49 (0.39, 0.62) | < 0.001 | 0.00 |
|  | Degree | 0.39 (0.29, 0.50) | < 0.001 | 0.00 |
| **Maternal Class** | V | *Reference* |  |  |
|  | IV | 1.24 (0.82, 1.86) | 0.307 | 22.42 |
|  | III Manual | 0.92 (0.61, 1.39) | 0.688 | 33.23 |
|  | III Non-manual | 0.84 (0.58, 1.21) | 0.331 | 27.36 |
|  | II | 0.85 (0.58, 1.21) | 0.387 | 29.05 |
|  | I | 0.62 (0.40, 0.95) | 0.025 | 3.65 |
| **Focus@9 Clinic Measures** | Adiponectin | 1.08 (1.01, 1.16) | 0.024 | 75.62 |
|  | BMI | 1.04 (1.00, 1.08) | 0.079 | 99.95 |
|  | IL-6 | 1.00 (0.94, 1.07) | 0.889 | 99.63 |
|  | Random Insulin | 0.99 (0.93, 1.06) | 0.817 | 99.56 |
|  | Leptin | 0.96 (0.86, 1.07) | 0.497 | 86.60 |
